# BAFF-R and CD21 dysregulation inhibits memory B cell persistence in patients with common variable immunodeficiency

**DOI:** 10.1101/2025.05.20.25327919

**Authors:** Mike Bogetofte Barnkob, Emilie Stavnsbjerg Larsen, Christine Nilsson, Zelvera Usheva, Caroline Louise Stougaard, Nina Bang, Emil Birch Christensen, Kim Martin Rasmussen, Line Lundegård Bang, Ditte Rask Tornby, Dorte K Holm, Julie Therese Skaugen, Christian Nielsen, Olivia Lie-Andersen, Sune Justesen, Lars Rønn Olsen, Rune Micha Pedersen, Thomas Emil Andersen, Torben Barington, Line Dahlerup Rasmussen

## Abstract

**Background:** Individuals with common variable immunodeficiency (CVID) are at increased risk of respiratory infections such as SARS-CoV-2 infection due to poorly understood defects within the memory B cell (MBC) compartment. The COVID-19 pandemic presented a unique opportunity to investigate the effects of a novel pathogen and vaccination on the immune system of patients with CVID.

**Methods:** A cross-sectional, single-center, cohort study was used to evaluate the immunologic effect of SARS-CoV-2 vaccination and/or disease on the immune system. We examined the antibody levels, neutralization capacity, MBC, and CD4^+^ T follicular helper cell (T_FH_) response against two SARS-CoV-2 variants in 23 CVID patients and 51 age- and sex-matched healthy individuals with both vaccine-induced and hybrid immunity across multiple immunization events.

**Results:** Our study shows that while CVID patients mount a sufficient CD4^+^ T_FH_ response against both ancestral and Omicron variants, and some levels of neutralizing antibodies, Spike-specific MBC formation is severely inhibited. Only CVID patients with hybrid immunity were able to generate switched MBCs against both variants. These MBCs were transient however, as Spike-specific switched MBCs did not persist over time in CVID patients. We found that the level of switched MBCs correlated with both CD21 and BAFF-R expression in patients, many of whom expressed low levels of either receptor.

**Conclusion:** Taken together, our study shows that CVID patients generate a sufficient CD4^+^ T_FH_ response against SARS-CoV-2 but rarely cross-reactive MBCs, and suggest that this effect is correlated with CD21 and BAFF-R dysregulation which might cause switched MBC obsolescence.

**Trial registration:** 21/54057.

**Funding:** Lundbeck Foundation, Novo Nordisk Foundation and Odense University Hospital.

## Introduction

Common variable immunodeficiency (CVID) is the most frequent symptomatic primary immunodeficiency among adults. CVID is characterized by low immunoglobulin concentrations, poor vaccine response and/or low numbers of switched memory B cells. The clinical picture is diverse, including increased susceptibility to respiratory pathogens such as severe acute respiratory syndrome coronavirus 2 (SARS-CoV-2) (1–3). The pathogenesis is believed to be a result of dysfunction within the memory B cell (MBC) compartment (4), although impairment of T cell helper function, including CD4+ T follicular helper (T_FH_) cells, can also play a role (5–7). With the introduction of novel vaccines during the corona virus disease (COVID-19) pandemic, a unique possibility occurred, in which effects of a novel pathogen and vaccines on the immune system could be explored in CVID patients.

Immunological memory forms the basis of protection against viruses following vaccination and infection. In healthy individuals, SARS-CoV-2 infection and mRNA vaccination stimulate antigen-specific B cells to differentiate in the germinal centers (GCs) of secondary lymphoid organs through instruction from helper CD4^+^ T cells, including T_FH_ cells (8,9). T_FH_ cells are specialised CD4^+^ T cells which provide important costimulatory signals and cytokines to B cells, promoting class switching, somatic hypermutation, and affinity maturation, ultimately leading to the generation of memory B cells (10,11). It has been shown that the CXCR5^+^ subset of CD4+ T cells (named circulating CD4^+^ T_FH_ cells) in peripheral blood share functional properties with bona fide T_FH_ cells, and can thus be studied as representative of the lymphoid T_FH_ cells. For SARS-CoV-2, these circulating T_FH_ cells have been found to correlate with spike-antibody production (12,13).

B cells that differentiate in the GC undergo affinity maturation through somatic hypermutation of the B cell receptor, becoming either long-lived plasma cells or MBCs (14). The former cells generate neutralizing antibodies, which wane over 6-12 months following an immunization event (15). In healthy individuals, MBCs persist long-term and play a crucial role in providing immunity against SARS-CoV-2 by allowing for rapid production of antibodies following re-exposure (16,17). In vaccinated individuals, this response is directed towards the virus receptor-binding domain Spike, which is also an immunodominant target following infection (18). Because new SARS-CoV-2 variants can rapidly acquire mutations in Spike, and antibody plasma concentrations decline, the effects of neutralizing antibodies diminish over time. However, both memory B and T cells persist long-term and retain protective functions against severe disease (19–21). MBCs in particular have been shown to encode a broad repertoire which allows protection against variants of the initial pathogen following antigen re-encounter (22–24). Interestingly, new immunization events against SARS-CoV-2, through either vaccine boosting or infection following the primary vaccination series, have been shown to increase the breadth and potency of MBC in healthy individuals (25–28).

In CVID patients, studies have primarily examined the antibody response following mRNA vaccination with either Moderna mRNA-1273 or Pfizer/BioNTech BNT162b2. These studies show that while patients are able to mount an antibody response against Spike following vaccination, especially after three or four doses (29), this response is severely blunted compared with healthy individuals (30–34). Few studies have closely examined the Spike-specific B cell memory profile (35,36) or Spike-specific CD4+ T_FH_ cells in CVID patients. In studies following either second or third vaccination, CVID patients had significantly lower frequencies of Spike-specific MBCs compared with healthy controls (37–39). It has also been shown that CVID patients with low frequency of switched MBCs and low-expression of CD21 on B cells have a poor humoral response to SARS-CoV-2 vaccination (30,32). It is currently unclear what effect repeated immunization events, including both infections and vaccinations, have on the antibody level and neutralization capacity among CVID patients, and whether novel viral variants can circumvent already acquired B-cell memory.

Here we performed a cross-sectional, single centre cohort study to examine the memory immune response in CVID patients using high-dimensional spectral flow cytometry, T-cell stimulation and microneutralization assays to analyze Spike-specific T_FH_- and B cells and their phenotype. In order to examine the robustness of these responses against unmet variants, we compiled a detailed vaccination and infection profile for each individual and analyzed their response against two variants of SARS-CoV-2, wild-type (WT, ancestral strain) and a BA.2 Omicron subvariant. This allowed us to closely examine both vaccine-only and hybrid immunity and how it affected B-cell memory formation and ability to cross-react to another variant. While Spike-specific T_FH_-cell responses were similar to healthy controls, CVID patients mounted poor antibody responses and had severely diminished Spike-specific MBC persistence, as these disappeared following immunization events. This effect was correlated with the surface expression of CD21 and B-cell-activating factor receptor (BAFF-R) on MBCs, underscoring these receptors’ importance in their persistence.

## Results

### Study participants and immunization profile

In total, 27 CVID patients and 51 healthy controls were included and had blood samples collected for further analysis (**Figure 1A**). Two CVID patients were neither vaccinated nor had any history of infection, and were not further examined. Among the remaining CVID patients, 23 (92.0%) fulfilled the revised ESID criteria, while 2 (8.0%) did not due to normal IgA levels or prior B-cell depleting therapy (**Table 1**).

**Figure 1.**
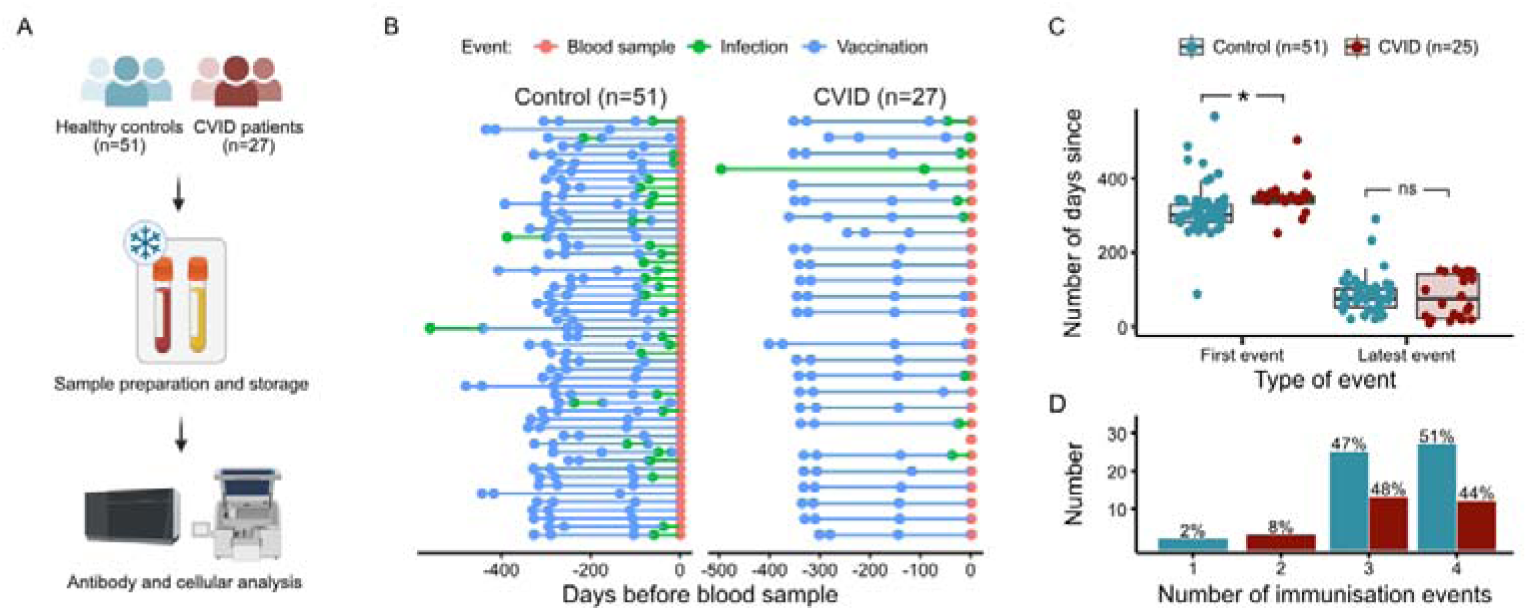
Study cohort and immunization events. A, Schematic of experimental setup used in this study. Initially, 27 CVID patients were included. Two patients were subsequently found not to fulfill ESID criteria, and further two did not experience any immunization events. **B,** Individual vaccinations (blue dots) and PCR-confirmed SARS-CoV-2 infections (green dots) for both groups up till time of blood sample (red dots). **C,** Time since first and latest immunization event in individuals with registered events (n=51 for healthy controls and 25 for CVID patients). **D,** Number and frequency of individuals per immunization events for healthy control (blue bars) and CVID patients (red bars). Abbreviations: CVID, common variable immunodeficiency disease. Boxplots correspond to median and 25th and 75th percentiles with whiskers ± IQR x 1.5. * = P < 0.05, by two-way ANOVA (C). Each dot (C) represents one individual.

**Table 1.** Baseline demographics of the cohort. Abbreviations: CVID, common variable immunodeficiency. IgRT, immunoglobulin replacement therapy. mAb, monoclonal antibody therapy. n, number. NA, not applicable. IQR, interquartile range. SARS-CoV-2, severe acute respiratory syndrome coronavirus 2.

| Demographics | CVID | Controls |
| --- | --- | --- |
| Number | 25 | 51 |
| Male: n (%) | 11 (44.0) | 25 (49.0) |
| Age in years: Median (IQR) | 55.5 (46.2-69.2) | 52.8 (33.2-66.1) |
| Age at CVID symptom onset: Median (IQR) | 36.1 (24.1-58.4) | NA |
| Age at CVID diagnosis: Median (IQR) | 46.8 (35.4-61.8) | NA |
| Born in Denmark: n (%) | 25 (100) | 51 (100) |
| Caucasian: n (%) | 25 (100) | 51 (100) |
| Met revised ESID CVID criteria: n (%) | 23 (92.0) | NA |
| Met old ESID CVID criteria (no IgA deficiency): n (%) | 1 (4.0) | NA |
| Revised ESID criteria not met due to prior Rituximab therapy: n (%) | 1 (4.0) | NA |
| Latest IgG level (g/L): Median (IQR) | 5.9 (4.7-8.3) | NA |
| **Clinical Manifestations** |  |  |
| Only infections: n (%) | 11 (44.0) | NA |
| Non-infections only: n (%) | 6 (24.0) | NA |
| Combined phenotype: n (%) | 8 (32.0) | NA |
| Immunoglobulin replacement therapy (IgRT) | 19 (76.0) | NA |
| **Type of IgRT** |  |  |
| Hizentra, s.c.: n (%) | 11 (57.9) | NA |
| HyQvia, s.c.: n (%) | 4 (21.1) | NA |
| Privigen, i.v.: n (%) | 4 (21.1) | NA |
| **Immunomodulating therapy** |  |  |
| Current systemic glucocorticoids: n (%) | 2 (8.0) | NA |
| Current anti-CD20 mAb: n (%) | 0 (0) | NA |
| Previous anti-CD20 mAb: n (%) | 3 (12.0) | NA |
| **SARS-CoV-2 vaccination regimes** |  |  |
| Pfizer + Pfizer | 1 (4.0) | 2 (3.92) |
| Pfizer + Pfizer + Pfizer | 8 (32.0) | 37 (72.55) |
| Pfizer + Pfizer + Pfizer + Pfizer | 13 (52.0) | 0 |
| Moderna + Moderna | 0 | 2 (3.92) |
| Moderna + Moderna + Moderna | 1 (4.0) | 5 (9.80) |
| AstraZeneca + Moderna + Moderna | 0 | 2 (3.92) |
| AstraZeneca + Moderna + Moderna + Moderna | 1 (4.0) | 0 |
| Johnson & Johnson + Moderna + Moderna | 0 | 2 (3.92) |
| No vaccinations | 1 (4.0) | 1 (1.96) |
| **Monoclonal Antibodies, n (%)** |  |  |
| Treated after project blood sample | 2 (8.0) | NA |
| Treated < 45 days before blood sample | 4 (16.0) | NA |
| Treated ≥ 45 days before blood sample | 2 (8.0) | NA |

The median age was 55.5 years (IQR: 46.2-69.2) for CVID patients and 52.8 (IQR: 33.2-66.1) for controls, and 44.0% of CVID patients and 49.0% of controls were male. For the CVID patients, the median IgG level was 5.9 g/L (IQR 4.7-8.3) and 76.0% received immunoglobulin replacement therapy (IgRT). Two patients (8.0%) were treated with low-dose oral glucocorticoids and 3 (12.0%) had received Rituximab after the CVID diagnosis. Monoclonal antibodies against SARS-CoV-2 (mAb; Sotrovimab) were given to 6 (24.0%) CVID patients constituting 54.5% of CVID patients with SARS-CoV-2 infections. The median time from mAb to study blood samples was 33 days. No controls had received mAb against SARS-CoV-2. Further characteristics are summarized in **Table 1**.

In **Figure 1B**, we show the immunization history for all participants. All participants with infection events were found to be anti-N positive, while all but one with no registered SARS-CoV-2 infection, were anti-N negative, indicating high accuracy of classification. The median time since the first immunization event was 344 days for CVID patients (IQR: 333-353 days), and 301 days for controls (IQR: 281-328 days). At study inclusion, 92.0% of CVID patients and 84.3% of controls had received three vaccine doses and 24.0% of CVID patients and 0% of controls had received four doses. The vaccination regimes are summarized in **Table 1**.

We decided to count both infection and vaccination as immunization events, and compare groups with same numbers of total immunization events. For both groups, the median time since the latest immunization event was 75 days (IQR: 51-101 days for controls, 21-142 for CVID patients) (**Figure 1C**). The majority of participants had three or four immunization events (**Figure 1D**).

### Antibody level and neutralization against SARS-CoV-2 WT and Omicron depend on the number of, and time since, the last immunization event

To examine the effect of immunization events on the antibody level and the degree of neutralizing capacity against SARS-CoV-2, we characterised the antibody response in CVID patients (n=23) and healthy controls (n=50). First, we compared the anti-Spike IgG levels after three and four immunization events. While both CVID patients and controls seroconverted, controls had significantly higher antibody levels than CVID patients, with approximately 12-fold and 5-fold higher antibody levels after three and four immunization events, respectively. Four immunization events resulted in a significant higher antibody concentrations than three in both groups, although the increase was larger for the control group (median: 35,602 BAU/mL, IQR: 17,889-40,000 BAU/mL) compared with the CVID group (median: 7,478 BAU/mL, IQR: 2,665-28,724 BAU/mL) (**Figure 2A**).

**Figure 2.**
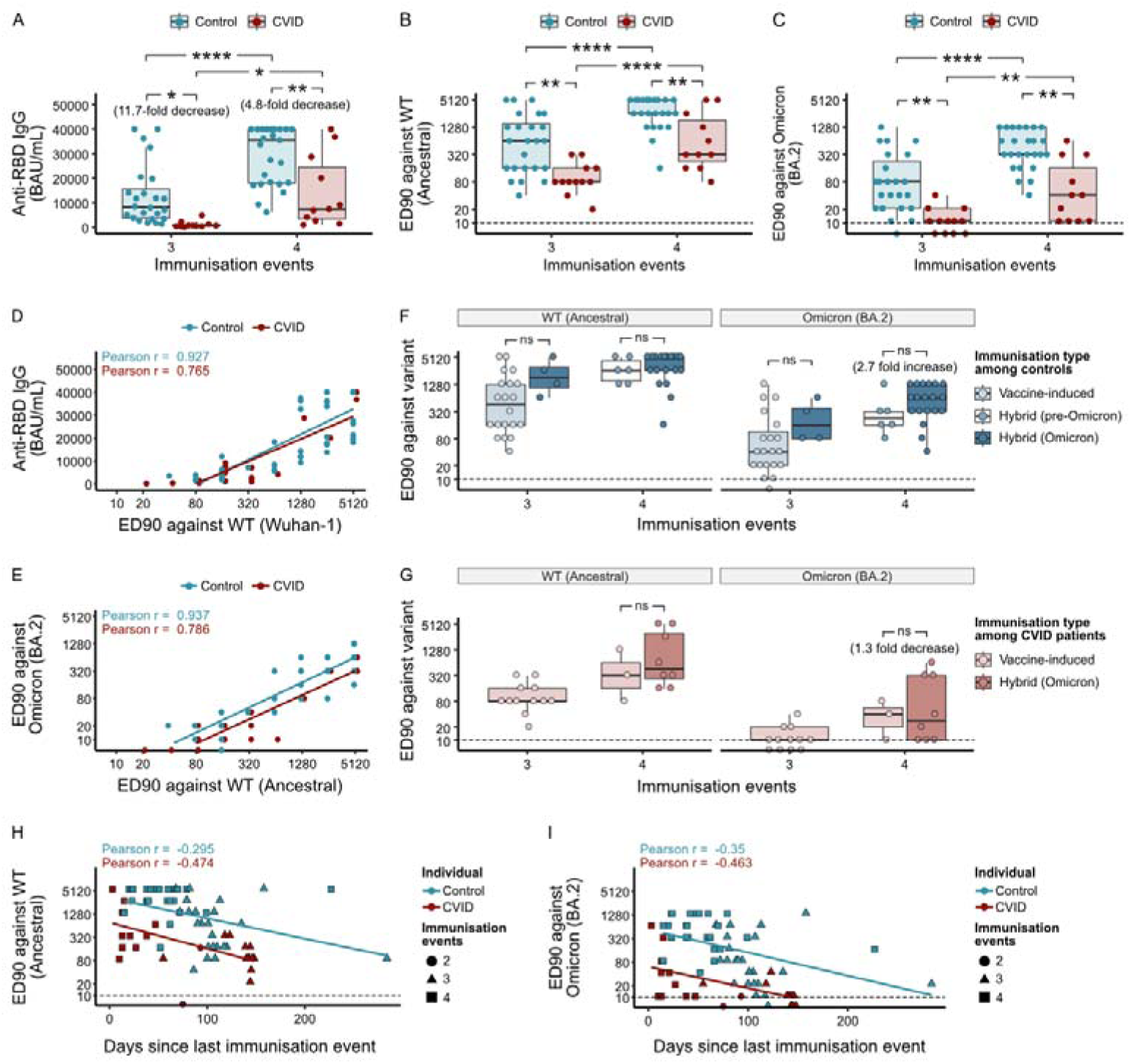
Antibody level and neutralization against SARS-CoV-2 WT and Omicron depend on the number of, and time since, the last immunization event. A, Anti-Spike RBD IgG antibody levels and immunization events. **B-C,** ED90 against WT (**B**) or Omicron BA.2 SARS-CoV-2 using serum from control or CVID patients. **D**, Correlation between Anti-Spike RBD IgG antibody levels and ED90 against WT Sars-Cov-2. **E,** Correlation between ED90 against WT and Omicron BA.2 SARS-CoV-2. **F-G,** ED90 against WT (left graph) or Omicron (right graph) per immunization event for control (**F**) and CVID patients (**G**). **H-I,** Correlation between ED90 against WT (**H**) or Omicron BA.2 (**I**) and time since last immunization event. Abbreviations: CVID, common variable immunodeficiency disease. ED90, effective dilution titre 90%. RBD, receptor binding domain. Boxplots correspond to median and 25th and 75th percentiles with whiskers ± IQR x 1.5. Lines (**D-E, H-I**) correspond to linear regression models for specified variables. Dash lines (**B-C, F-I**) correspond to the threshold level of neutralization. * = P < 0.05, ** = P < 0.01, **** = P < 0.0001 by two-way ANOVA (**A-C, F-G**). Each dot represents one individual.

To qualify antibody functionality in each group, we next investigated serum neutralization against live authentic virus for both WT (**Figure 2B**) and Omicron BA.2 (**Figure 2C**) in a microneutralization assay (40). Briefly, we quantified the highest dilution protecting 90-100% of Vero-E6 cells from viral induced cytopathic effect, here called effective dilution titre (ED90). We have previously shown that anti-Spike IgG levels in IgRT products at this point in the SARS-CoV-2 pandemic were negligible (29) thus allowing us to compare virus neutralization between CVID patients and controls. We found that anti-Spike IgG levels correlated closely with neutralization activity (**Figure 2D**). Virus neutralization was more potent in the control group, across immunization events and against both variants. While all patients had sufficient neutralization capacity against WT virus, 39% of CVID patients (9/23) had insufficient response against Omicron, compared to 6% (3/50) among controls (**Figure 2C**). After four immunization events, CVID patients were significantly better at neutralizing both WT and the Omicron variant than patients with only three events (**Figure 2B-C**). Neutralization of WT correlated closely to Omicron BA.2 neutralization for both groups (**Figure 2E**).

Next, we wanted to estimate whether the neutralization activity differed according to hybrid immunity and vaccine only-immunity. We compared vaccine-only immunity with hybrid immunity, arising from either pre-Omicron or Omicron infections after vaccination (**Figure 2F-G**). No significant difference among immunity profiles between individuals with the same number of immunization events was found. However, the sample size of the CVID group was small.

Finally, we examined the waning of the humoral response over time by comparing the time since the last immunization event with neutralization activity against WT (**Figure 2H**) and Omicron BA.2 (**Figure 2I**). CVID patients had poorer neutralization following immunization (intercept titer against WT and Omicron was 1:1631 and 1:158, respectively, at day 0) than healthy controls (intercept titer against WT and Omicron was 1:3367 and 1:659, respectively, at day 0). We next calculated the decay-rate and half-life of the neutralization response over time. Both groups experienced a waning over time (**Figure 2H-I**). For controls, the neutralization response was halved in 57.4 and 51.1 days against WT and Omicron BA.2, respectively, while CVID patients had a half-life of 44.6 and 51.1 days.

### CVID patients retain few switched memory B cells against SARS-CoV-2

We next examined the B cell compartment by multiparametric flow cytometry (**Supplementary** Figure 1). We found no differences in overall B cell frequencies (**Figure 3A**), but as expected CVID patients had substantially fewer switched MBCs (**Figure 3B-C**). No differences were found in overall MBCs or plasmablast frequencies (**Figure 3C**).

**Figure 3.**
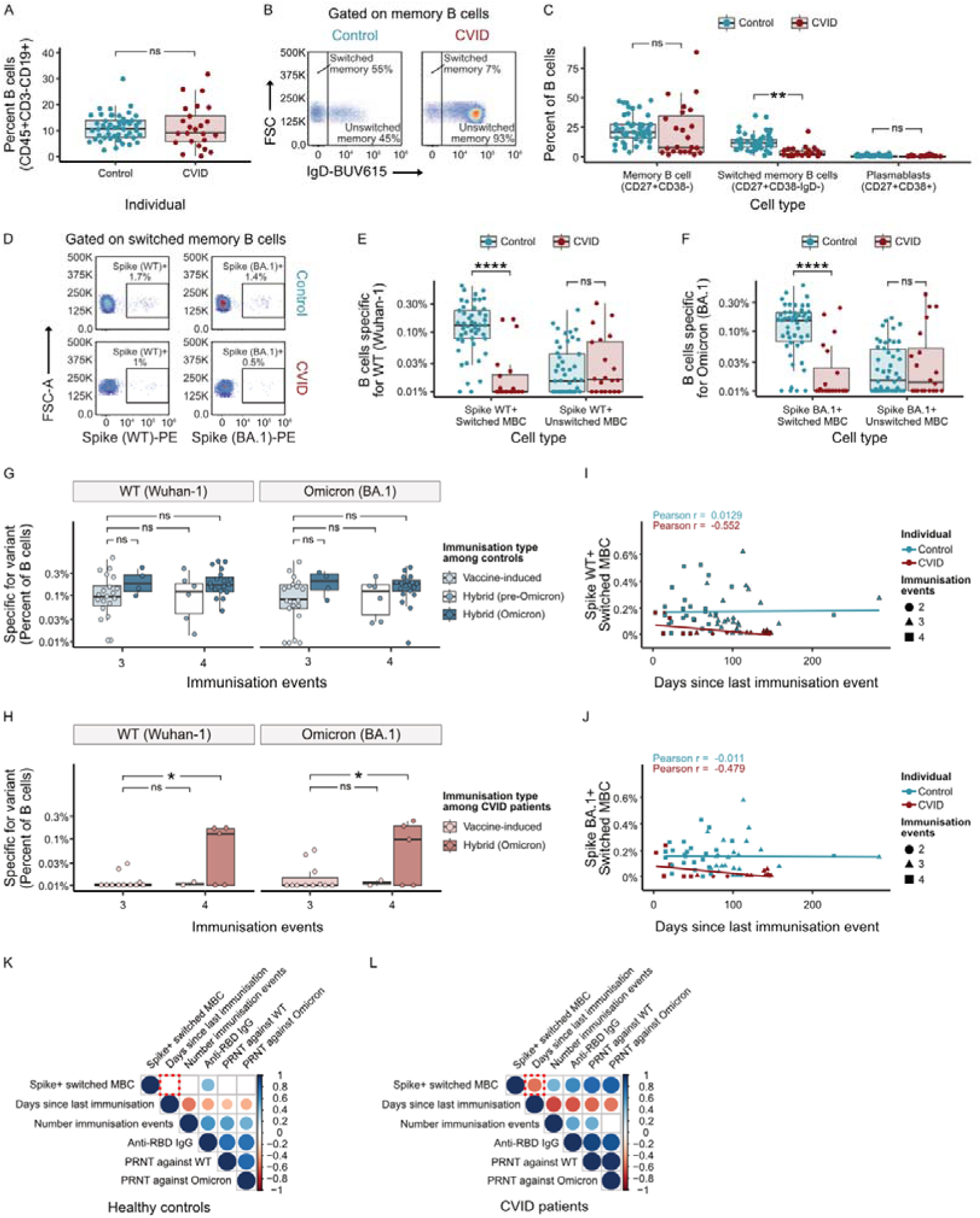
CVID patients retain few switched memory B cells against SARS-CoV-2. A, Percentage of B cells in control and CVID patients. **B,** Representative gating showing frequencies of switched and unswitched memory B cells (following gating onto memory B cells, see supplementary Figure 1) in controls and CVID patients. **C,** Frequencies of MBCs, switched MBCs, and plasmablasts. **D,** Representative gating showing staining of switched MBCs specific for recombinant Spike WT or Spike Omicron (BA.1 variant). **E-F**, Frequencies of switched and unswitched MBCs specific for Spike WT (**E**) or Omicron (**F**). **G-H,** Frequency of MBCs specific for Spike WT (**G**) or Omicron (**H**) by immunization history. **I-J**, Frequency of Spike-specific MBCs plotted against days since last immunization event. **K-L,** Correlation matrix comparing key variables measured concerning antibody level, immunization history and neutralization against SARS-CoV-2 in healthy controls (**K**) and CVID patients (**L**). Red dashed box indicates primary variable between control and CVID patient, showing a negative correlation with Spike-specific switched MBCs in CVID patients. Abbreviations: CVID, common variable immunodeficiency disease. ED90, effective dilution titre 90%. MBC, memory B cell. Boxplots correspond to median and 25th and 75th percentiles with whiskers ± IQR x 1.5. Lines (**I-J**) correspond to linear regression models for specified variables. * = P < 0.05, ** = P < 0.01, **** = P < 0.0001 by student’s T test (**A**) or two-way ANOVA (**C, E-H**). Each dot represents one individual.

In order to identify Spike-specific MBCs, we included recombinant Spike WT or Omicron (BA.1) protein in our panel, allowing us to see which B cells, through B-cell-receptor (BCR) binding to the proteins, were specific for either variant (**Figure 3D**). In both healthy controls and CVID patients, a large proportion of individuals had very low or undetectable levels of unswitched MBCs specific for either Spike variant and no difference in frequencies was found. However, for switched MBCs, almost no CVID patient had any detectable clones, whereas almost all healthy individuals had (**Figure 3E-F**).

We then examined the effects of immunization history on the level MBCs specific for either variant. We found that healthy controls with vaccine-induced immunity generated similar frequencies of Spike-specific MBCs as controls with hybrid immunity followed by an Omicron infection. This was the case against both Spike WT and against Spike Omicron, a variant that vaccine-only individuals had not seen. Frequencies were similar after both three and four immunization events (**Figure 3G**). However, for CVID patients, significant levels of Spike-specific MBCs against WT or Omicron Spike were only seen in the few individuals with hybrid immunity (**Figure 3H**).

We next examined the persistence of MBCs. Whereas MBCs specific for either WT or Omicron Spike could be found up to and beyond 200 days following an immunization event in healthy controls, almost no one could be found in CVID patients after 30 days (**Figure 3I-J**). Fitting a linear regression model to the frequency of MBCs specific for Spike WT over time, did not reveal any dependence of time for healthy controls (Pearson’s r = 0.01), whereas time was negatively correlated with MBCs in CVID patients (Pearson’s r = −0.55) (**Figure 3I**). Time since last immunization was negatively correlated with switched MBCs across a number of correlations associated with antibody level, neutralization (ED90) and immunization events in CVID patients, but not in healthy controls (**Figure 3K-L**).

### BAFF-R and CD21 dysregulation affect switched memory B cells frequencies in CVID patients

As our analysis suggested that CVID patients can generate Spike-specific switched MBCs, but not retain them, we next wanted to examine the B-cell compartment in more detail. We first evaluated the overall B cell profile of our cohort by using uniform manifold approximation and projection (UMAP) where subsets were classified using standard B-cell markers (41,42) (**Figure 4A**). As the UMAP indicated less diverse subpopulations within the B-cell compartment of CVID patients (**Figure 4B**), especially within MBCs, we calculated Shannon diversity index for each cohort. This analysis takes into account both the number of cells and the relative abundance of each subpopulation and allows us to compare the diversity of subpopulations between groups. Our analysis showed that CVID patients were significantly less diverse in their B-cell subpopulations than controls (**Supplementary** Figure 2A). When examining the relative frequency of B-cell subsets across individuals, it was clear that CVID patients had highly individual derangements across a number of different B-cell subsets (**Figure 4C**), underscoring the heterogeneity of the disease.

**Figure 4.**
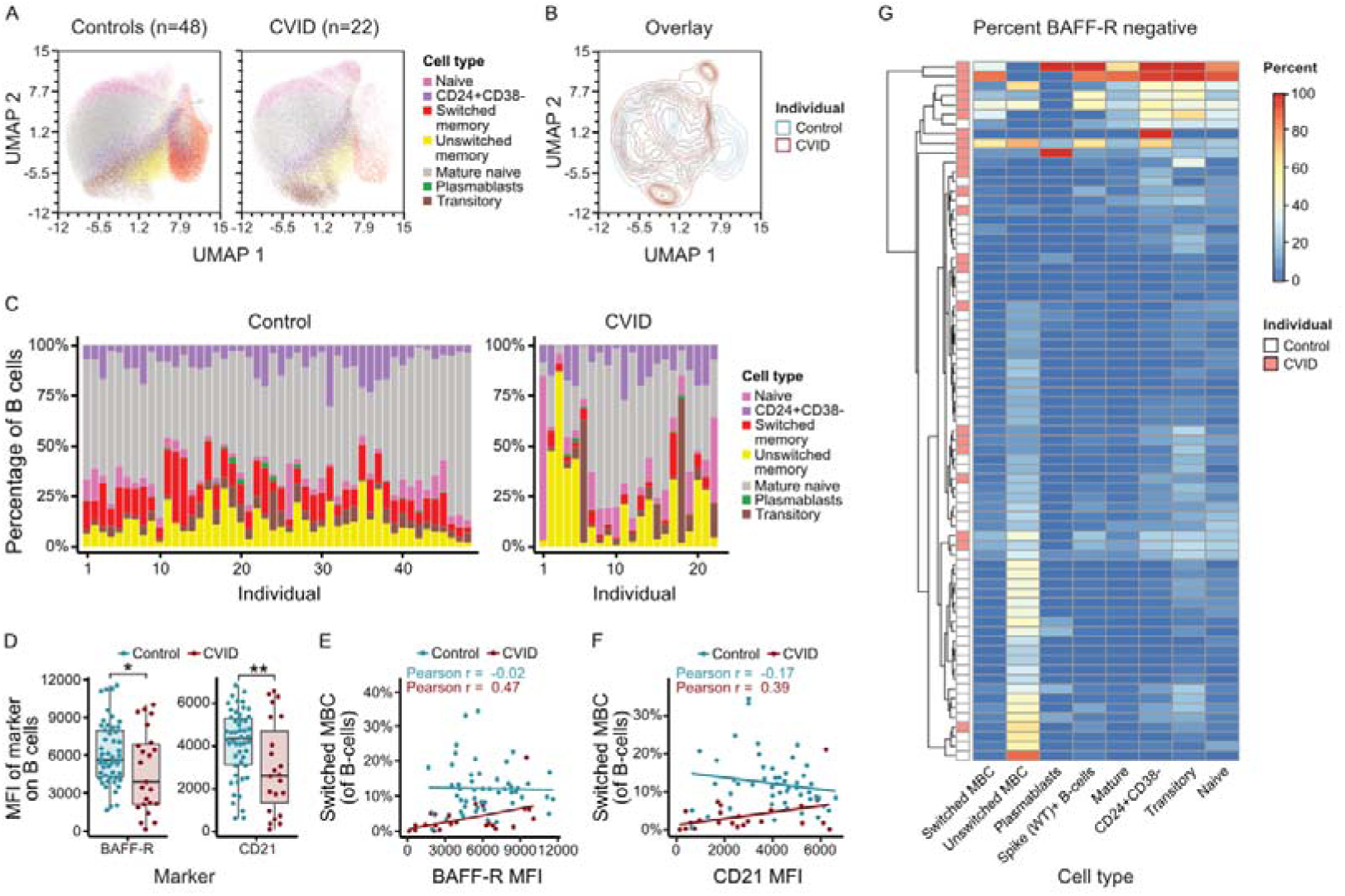
Low BAFF-R and CD21 expression correlates with low levels of switched memory B cells. A, UMAP of circulating B-cell compartment from control (left, n=48) and CVID (right, n=22) with canonical B cell subsets marked (gating shown in Supplementary Figure 1). **B**, Overlaid contour plot of UMAPs from control (blue) and CVID (red) individuals. **C**, Relative frequencies of B cell subsets from individual control (left) and CVID (left) patients. **D**, MFI of BAFF-R (left) and CD21 (right) on all B cells among control and CVID patients. **E-F**, Frequency of switched MBCs plotted against MFI of BAFF-R (**E**) and CD21 (**F**). **G**, Dendrogram showing percentage of B cells negative for BAFF-R across subsets, grouped by hierarchical clusterings. Individuals marked by white (control) or red box (CVID patient). Boxplots correspond to median and 25th and 75th percentiles with whiskers ± IQR x 1.5. Abbreviations: MBC, memory B cell. MFI, mean fluorescence intensity. UMAP, uniform manifold approximation and projection. Lines (**E-F**) correspond to linear regression models for specified variables. * = P < 0.05, ** = P < 0.01 by Student’s t test (**D**). Each dot represents one individual.

We wanted to understand what might drive this difference between controls and CVID patients. Across all B-cell markers measured, the receptors BAFF-R and CD21 were differentially expressed between controls and CVID patients (**Figure 4D**). Both receptors are involved in MBC survival and persistence and genetic defects in *TNFRSF13C* (encoding BAFF-R) and *CR2* (encoding CD21) have been associated with CVID (43,44). However, none of the patients in our cohort had clinical relevant variants in either gene, showing that protein expression was affected at another stage. BAFF-R has previously been shown to be down-regulated due to cleavage of the receptor following BAFF binding (45). Indeed, when we cultured B cells from healthy donors on feeder cells expressing either CD40 Ligand (CD40Lg) alone or both CD40Lg and BAFF, we noticed almost complete loss of BAFF-R on the cell-surface of B cells when BAFF was present (**Supplementary** Figure 2B-C).

For both receptors, fewer molecules per cell on all B cells corresponded to fewer overall switched MBCs (**Figure 4E-F**). Likewise, the expression of both receptors correlated positively with the frequency of switched MBCs in CVID patients (Pearson’s r = 0.47 and 0.39 for BAFF-R and CD21, respectively), but not in controls (Pearson’s r = −0.02 (p-value = 0.87) and −0.17 (p-value = 0.24) for BAFF-R and CD21, respectively) (**Figure 4E-F**). More broadly, BAFF-R negative cells were found across B-cell subsets, correlating with a cluster of CVID patients (**Figure 4G**). Both BAFF-R and CD21 dysregulation are thus associated with the formation or persistence of MBC cells.

### CVID patients generate antigen-specific circulating CD4+ T_FH_ cells

In order to examine whether the diminished frequency of switched MBCs in CVID patients was due to a lack of support from antigen-specific CD4^+^ T_FH_ cells, we stimulated PBMCs with peptide pools specific for either WT Spike or Omicron BA.2 Spike. Cells were also stimulated with CD3/CD28 beads, which confirmed that all patients in this cohort could raise a relevant T cell response. We then examined activation induced markers (AIM) on non-naïve CD4^+^ T cells (see gating in **Supplementary** Figure 3) by flow cytometry as defined by CD69 expression and IFNγ and/or TNF expression (**Figure 5A**). We found no difference in the frequency of circulating T_FH_ cells able to react to either WT or Omicron BA.2 Spike peptides between CVID patients or controls (**Figure 5B**).

**Figure 5.**
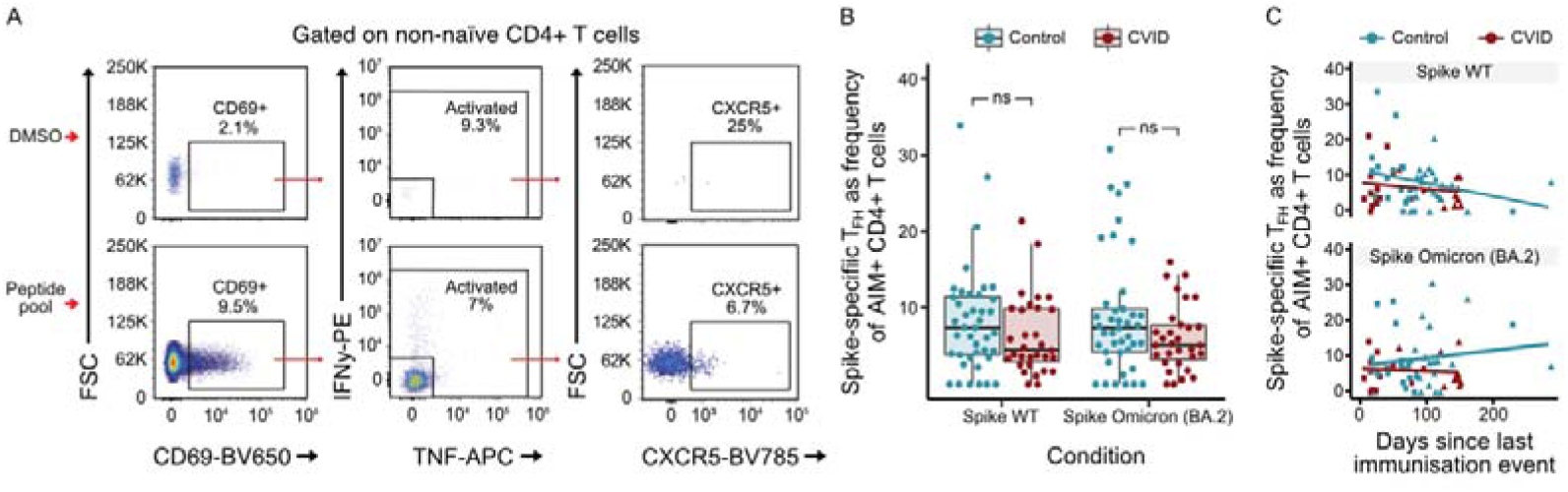
Analysis of peptide-inducible Spike-specific circulating CD4^+^ T_FH_ cells. A, Gating strategy to identify AIM^+^CD4^+^ T_FH_ cells (following gating onto non-naïve CD4^+^ T cells, see supplementary Figure 2). **B,** Frequency of AIM^+^CD4^+^ T cells which are circulating T_FH_ cells against Spike WT or Omicron (BA.2) peptide pools. Boxplots correspond to median and 25th and 75th percentiles with whiskers ± IQR x 1.5. **C,** Frequency of AIM^+^CD4^+^ T cells which are circulating T_FH_ cells plotted against days since last immunization event. Lines (**C**) correspond to linear regression model for specified variables. ns = not significant by Mann-Whitney test (**B**). Each dot represents one individual.

Circulating T_FH_ have been detected up to 6 months following immunization against SARS-CoV-2 (46). We therefore correlated the frequency of circulating T_FH_ cells with the number of days since the last immunization event. For CVID patients the correlation coefficient against WT peptides or Omicron BA.2 specific-peptides indicated no correlation (Pearson r = 0.-19 (p-value = 0.35) and −0.1 (p-value = 0.64) against Spike WT and Omicron BA.2, respectively) (**Figure 5C**). Taken together these data indicate that the majority of CVID patients generated a sufficient antigen-specific CD4+ T_FH_ response which lasts at least 6 months and is comparable to healthy controls.

## Discussion

In this study, we characterized the B-cell memory immune response against SARS-CoV-2 in CVID patients across diverse immunization events (**Figure 1**). A major strength of the study was the access to high quality data concerning infection and vaccination status for both the CVID cohort and an age-and sex matched healthy cohort from the general population in a country with a high SARS-CoV-2 test coverage. This allowed for a detailed immunization history to be made for each participant of the study. By combining functional assays with high-dimensional flow cytometry and the ability to identify Spike-specific MBCs, we were able to thoroughly examine the B-cell response to SARS-CoV-2 variants of concern. Our analysis revealed profound deficits in humoral immunity and memory B-cell persistence. Importantly, we were able to take into account multiple immunization events, including both vaccinations and infections into our analysis, and test cross-reactivity to a variant of concern. Our findings underscore the poor vaccine responsiveness in CVID, and indicate an association between BAFF-R and CD21 dysregulation and MBC development.

Consistent with previous reports (30–34), CVID patients in our cohort demonstrated significantly lower anti-Spike IgG levels and weaker neutralizing antibody responses against both the ancestral SARS-CoV-2 strain and the Omicron BA.2 variant compared to healthy controls (**Figure 2**). Although we observed enhanced antibody titers and neutralization capacity in CVID patients with repeated immunization events, the magnitude of this response remained reduced compared to healthy controls. Using linear regression analysis, we found that the titer at day 0 was lower for CVID patients, suggesting an inability to produce sufficient quantities of neutralizing antibodies directly following an immunization event. However, the half-life of neutralization activity of antibodies was comparable between CVID patients and controls. Taken together these data indicate that the primary limitation in CVID is due to the quality of antibodies produced by B cells or the number of antibody-producing cells that can be stimulated.

In line with these observations, a key result of our study is the failure of CVID patients to sustain long-term Spike-specific MBCs following SARS-CoV-2 immunization, across immunization events and type (**Figure 3**). Others have similarly found fewer MBCs following vaccination in CVID patients (35,38,39), but not taken into account infection histories. Here we find that while most healthy individuals exhibited robust and durable antigen-specific switched MBCs over time, including MBCs that could bind to a variant of Spike not yet experienced, CVID patients lacked detectable levels of these cells beyond 30 days post immunization or infection. Novel viral variants are thus expected to more easily circumvent acquired B-cell immunity in patients with CVID. Indeed, only those CVID patients who had recently encountered the virus or vaccine, had measurable Spike-specific switched MBCs. Still, immature unswitched Spike-specific MBCs could be found in CVID patients. These data thus highlight a fundamental defect at this stage in the development of B-cell memory against SARS-CoV-2.

Because CD4^+^ T_FH_ cells promote long-lived humoral immunity after vaccination by providing help to B cells within the GC (15), we examined whether the lack of switched MBCs were due to defects within this cell-population. However, we found no difference among circulating T_FH_ cells reactive for either SARS-CoV-2 WT or Omicron, for CVID patients compared to controls (**Figure 5**). Other groups have similarly examined this T-cell subset in CVID patients, and come to various conclusions. Our data is in line with Gupta et al. (47) and Steiner et al (39), but not Sauerwein et al. (37) who find an impaired CD4^+^ T_FH_ response following vaccination in a smaller cohort of patients. These differences could be due to differences in the number of immunization events or because we specifically examined non-naive CD4^+^ T cells.

Instead, this impaired MBC persistence was correlated with lower surface expression of BAFF-R and CD21 on B cells (**Figure 4**), receptors critical for B cell survival and homeostasis. Our data suggest that reduced expression of these receptors — despite the absence of pathogenic variants in *TNFRSF13C* or *CR2* in our cohort of CVID patients — contributes to the loss of B-cell diversity and memory longevity in CVID. While low CD21 expression has previously been shown to affect humoral immunity following SARS-CoV-2 vaccination (30,32), in agreement with our data, less is known about BAFF-R (48). Here we find that a large subset of CVID patients express low levels of the receptor across B-cell subsets.

BAFF-R is a key member of the TNF receptor superfamily, and plays an essential role in B cell biology by promoting survival, proliferation, and differentiation upon binding to its ligand BAFF (BlyS, TNFSF13B) (49,50). Signaling through BAFF-R activates both the non-canonical NF-κB pathway via NF-κB-inducing kinase (NIK) and Inhibitor of nuclear factor kappa-B kinase subunit alpha (IKK-α), and the canonical pathway via IKK2, supporting B cell homeostasis and function. In recent animal studies, BAFF-R signaling has been shown to be crucial not only for the viability of naive and mature B cells but also for the long-term maintenance of MBCs and the sustainability of GC responses (51,52). While BAFF-R defective mice are thus able to mount GC responses following immunization, these are not sustained (53,54), reminiscent of our results here.

Despite these insights from animal studies, the role of the BAFF/BAFF-R pathway in human MBCs is less clear. Evidence from systemic lupus erythematosus (SLE) patients treated with the BAFF-depleting antibody belimumab, indicate that BAFF-depletion does not affect existing MBCs, but does inhibit antibody response to subsequent seasonal influenza vaccinations (55). In CVID patients, most patients are found to have high BAFF levels (56–58), but low BAFF-R expression on both naive and MBCs (45). Both situations can thus possibly affect the pathway in similar ways. Although mutations in *TNFRSF13C* (encoding BAFF-R) have been identified in a small subset of CVID patients (43), <1% of patients harbor such mutations (59), suggesting alternative mechanisms of BAFF-R downregulation. Among CVID patients, down-regulation of BAFF-R can happen following chronic increase in BAFF, due to binding-induced cleavage of the receptor (45), which has also been described for patients with Sjögren’s syndrome and SLE (60). Indeed, we found that BAFF-R is down-regulated when B cells from healthy individuals are grown on feeder cells expressing both CD40L and BAFF, but not when only CD40L is present on feeder cells. Overall, these features suggest that disruption in BAFF/BAFF-R signaling negatively affects MBC survival and that this pathway could represent a potential pharmacological target.

The limitations of our study include the cohort size, for which e.g. the number of CVID patients with hybrid immunity was small, limiting the statistical power for subgroup analyses. Furthermore, while we assessed MBC frequency and phenotype, functional assays evaluating recall responses upon antigen re-exposure could further delineate the extent of immune memory impairment. Analysis of SARS-CoV-2 neutralization among patients could also be affected by mAb treatments, which six CVID patients received prior to analysis. Finally, longitudinal sampling beyond the time points analyzed here would be valuable to fully assess the kinetics of MBCs in CVID and increase causal inference.

In conclusion, our findings demonstrate that CVID patients exhibit a defect in the formation and persistence of Spike-specific switched MBCs following SARS-CoV-2 immunization, possibly linked to BAFF-R and CD21 dysregulation. These results provide initial mechanistic insight into the poor vaccine responsiveness in CVID by implicating the BAFF/BAFF-R pathway as important for MBCs persistence.

## Methods

### Human subjects

#### Sex as a biological variable

Participants from both biological sexes were included in the study. The cohorts were sex-matched, and included 11 (44%) and 25 (49%) males in the CVID and healthy control cohort, respectively. Sex was not considered as a biological variable.

#### Study approval

78 individuals (51 healthy controls and 27 CVID patients) provided oral and written informed consent and were enrolled in the study. The study was approved by The Regional Committees on Health Research Ethics for Southern Denmark (journal number 21/54057), by The Danish Data Protection Agency (journal number 21/60870), and performed according to the Declaration of Helsinki.

#### Setting

The prevalence of CVID in Denmark is 1:26,000 (61). The health care system in Denmark is tax-supported and immunoglobulin replacement therapy (IgRT), as well as vaccination, testing and treatment for SARS-CoV2 are provided free of charge. In the beginning of the pandemic, the test capacity was limited. However, after the first wave, Denmark used an extensive, nationwide testing program that included testing both symptomatic and non-symptomatic patients. CVID patients were among the first groups to be vaccinated.

#### Population

We included all CVID patients at Odense University Hospital (OUH), 18 years or older, who consented to the study. CVID was defined according to the revised European Society for Immunodeficiency (ESID) diagnostic criteria for CVID (62,63). For every CVID patient, two healthy age- and sex matched controls were recruited from the Danish Blood Donor corps at OUH.

#### Study design

We used a cross-sectional, single-center, cohort study to evaluate the immunologic effect of SARS-CoV2 vaccination and/or disease on the immune system for CVID patients compared to healthy controls.

#### Data collection

Based on a chart review, we extracted data concerning date of birth, sex, CVID diagnosis, clinical phenotype, results of blood tests, immunologic data, genetic information and use of medication. Data on time and result of SARS-CoV-2 testing, including variants, were retrieved from the Danish Microbiology Database. Data concerning type of vaccination was retrieved from the Danish vaccination registry. All CVID patients and controls had blood drawn for a biobank between January and March 2022.

#### Exposure

Immunization events were defined as a vaccination event or registration of a positive SARS-CoV-2 test independently of admission. Seroconversion was defined as anti-Spike IgG levels ≥7.1 BAU/mL. Neutralization against WT virus was defined as a titer > 1:10. At the time of the study, anti-Spike IgG levels in IgRT products had been shown negligible (29).

#### Genetic analysis

*TNFRSF13C* and *CR2* variants were interpreted on whole-genome sequencing done at time of CVID diagnosis by the Department of Clinical Immunology, OUH, and classified according to American College of Medical Genetics and Genomics guidelines (64).

### Sample processing

Venous blood was collected into sodium heparin and EDTA sample tubes by standard phlebotomy. Blood was diluted 1:1 with medium containing RPMI 1640 with GlutaMAX supplement (Gibco, catalog number 61870010), 10% heat inactivated fetal bovine serum (FBS), and 100 U/mL Penicillin, and 100 μg/mL Streptomycin (P/S) (here called R10). The sample was then layered onto 50 mL SepMate tubes (STEMCELL Technology, catalog: 85450) containing Ficoll-Paque PLUS (Cytiva, catalog: 17144002). Sample tubes were centrifuged at 1000 g for 20 minutes, and peripheral blood mononuclear cells (PBMCs) were harvested by sterile disposable pipette into a new 50 mL tube, which was filled with PBS (Gibco, catalog: 10010015) with 2% FBS, mixed and centrifuged at 500 g for 10 minutes. This washing step was repeated twice. Cell number and viability was determined. PBMCs were then cryopreserved in R10 with 10% dimethyl sulfoxide (DMSO) and stored at −80°C until analysis. Plasma samples were centrifuged and stored at −20°C until analysis.

### SARS-CoV-2 antibodies

All SARS-CoV-2 serological testing was conducted with serum. Anti-Spike IgG antibodies were determined using the Alinity SARS-CoV-2 IgG II Quant assay (Abbott Laboratories) according to the manufacturer’s specification. This assay provides a quantitative determination of IgG antibodies to the receptor binding domain (anti-RBD) of the S1 subunit of the spike protein of SARS-CoV-2, and is calibrated against the WHO International Standard for anti-SARS-CoV-2 immunoglobulin (NIBSC code 20/136). Results are provided as standardised binding antibody units (BAU)/mL. Arbitrary units (AU) from the assay were converted to BAU with the following equation: 1 BAU/mL = 0.142 × AU/mL. Levels ≥7.1 BAU/mL were considered positive and the individuals defined as responders.

SARS-CoV-2 nucleocapsid IgG antibodies (anti-N) were determined using the qualitative Alinity SARS-CoV-2 IgG assay (Abbott Laboratories).

### Microneutralization assay

In order to assess antibody efficiency against live virus, we used a microneutralization assay modified according to our previously published protocol (40). All work with authentic SARS-CoV-2 was performed in an approved Biosafety Level 3 (BSL-3) laboratory (license number 20200016905/5). Patient CPDA-plasma samples were diluted 1:8 in 37°C preheated complete viral culture medium (Dulbecco’s modified Eagle medium with 2% heat-inactivated fetal bovine serum, 1% penicillin, streptomycin and amphotericin B in a concentration of 2.5 mg/L). Samples were incubated for 30 minutes at 37°C on a shaker table to allow for the samples to clot completely. The sample supernatant was diluted in cold complete viral culture medium in two-fold serial dilutions (ranging from 1:8 – 1,024) and transferred to sterile 96-well microtitre plates. Authentic SARS-CoV-2 strains were diluted to approximately 2,000 plaque-forming unit (PFU) per mL in cold complete viral culture medium. Virus and plasma were mixed, yielding a final plasma dilution ranging from 1:10 to 1:1,280 and a final virus concentration of 200 PFU/mL corresponding to approximately 100 TCID50 (50% tissue culture infectious dose) per mL. Virus-plasma mixtures were incubated for 30 minutes at 37°C in a humidified atmosphere with 5% CO_2_ and 50 µL were subsequently transferred onto a confluent layer of Vero-E6 cells (ATCC, catalog: CRL-1586) in 96-well microtitre plates and incubated for another hour. One hundred µL of 37°C preheated complete viral culture medium were added to each well and the plates were incubated for 4 days at 37°C in a humidified atmosphere with 5% CO_2_. Virus-plasma mixture was removed and discarded and 50 µL of 10% neutral buffered formalin solution (Sigma-Aldrich) were added to each well and incubated for at least 20 min. Fixative was removed and cells were stained with 5% Gram’s crystal violet solution (Sigma-Aldrich) for 3-5 min. Wells were emptied and remains were removed by tapping the plate against a cloth. The effective dilution titre (ED90) was defined as the highest dilution protecting 90-100% of Vero-E6 cells from virally induced cytopathic effect (CPE). Patient samples were tested in duplicates along with two positive and two negative controls. We obtained ED90 titres towards clinical isolates of an ancestral and a BA.2 SARS-CoV-2 strain. Genomic sequences can be accessed at GenBank (accession number ON055855 and ON055857, for ancestral and BA.2 SARS-CoV-2 strains, respectively).

### Flow cytometry

#### Flow cytometry

Data was acquired on a SONY ID7000 spectral cytometer with five lasers (355/405/488/561/637 nm). Prior to acquisition, the instrument was calibrated using AlignCheck particles (Sony Biotechnology, catalog: AE700510) and the performance 8-peak beads (Sony Biotechnology, catalog: AE700522), following manufacturer’s guidelines. In each experiment, spectra were generated using single-stained samples consisting of antibodies in Ultracomp eBeads compensation beads (Thermo Fisher, catalog: 01-2222-41) for each individual antibody.

#### Detection of Spike-specific B cells

Tubes with PBMCs were thawed and resuspended in 5 mL of R10 with 10 IU ml^−1^ DNase I (Sigma-Aldrich, catalog: 11284932001), before being spun down at 500 g for 5 minutes. Cells were then washed in PBS with 1% bovine serum albumin (BSA) (500 g for 5 minutes), before being aliquoted to three wells per individual. In two of these wells, 200 ng recombinant SARS-CoV-2 Spike WT-biotin protein (Miltenyi Biotec, catalog: 130-127-681) or 200 ng recombinant SARS-CoV-2 Spike Omicron-biotin protein (Miltenyi Biotec, catalog: 130-130-417) were added in 100 uL of PBS with 1% BSA, while the final well only had PBS with 1% BSA added. Samples were left to incubate for 1 hour at 4°C, washed, and then stained or 30 minutes at 4°C in PBS with 1% BSA using the following antibodies f: CD45 (APC-H7), CD3 (BV480), CD14 (BV480), CD19 (BUV563), CD20 (BV750), IgD (BUV615), CD21 (V450), CD27 (BV711), CD24 (FITC), CD138 (PerCP-Cy5.5), CD38 (BB790), BAFF-R (APC), and Streptavidin-PE. Cells were then washed twice in PBS before analysis. Gating strategy is outlined in **Supplementary** Figure 1.

#### Activation induced marker (AIM) assay

Tubes with PBMCs were thawed and resuspended in 5 mL of R10 with 10 IU ml^−1^ DNase I (Sigma-Aldrich, catalog: 11284932001), before being spun down at 500 g for 5 minutes. Cells were then resuspended in R10 and approximately 10^6^ cells per well plated in a U-bottom 96 well plate and allowed to rest for 2-4 hours at 37°C. Cells from each individual were then stimulated with either a SARS-CoV-2 Spike WT peptide pool (PepTivator SARS-CoV-2 Prot_S B.1.1.529/BA.2 WT Reference Pool, Miltenyi Biotec, catalog: 130-130-806), Spike BA.2 peptide pool (PepTivator® SARS-CoV-2 Prot_S B.1.1.529/BA.2 Mutation Pool, Miltenyi Biotec, catalog: 130-130-807), CD3/CD28 beads (Gibco, catalog: 11132D) as a positive control or DMSO, as a negative control in a total volume of 200 uL per well. The Spike BA.2 peptide pool consisted of 68 peptides of 15 amino acids (aa), which cover the mutated regions compared to WT spike. The Spike WT pool covers the homologous domains of the Wuhan sequence. Approximately 120 pmol peptides were added to each well with peptide pools. Cells were left to incubate for 16 hours in a 37°C humidified incubator with 5% CO_2_. Cells were spun down and resuspended in R10 with Brefeldin A (BioLegend, catalog: 420601) and cells incubated for another 4 hours. Cells were then washed in PBS with 1% BSA (500 g for 5 minutes) and stained for the following surface markers: CD45 (BV570), CD14 (Pacific Blue), CD19 (Pacific Blue), CD3 (APC-Fire 750), CD4 (SparkBlue-550), CD8 (PE-Cy7) and CD69 (BV650) for 30 minutes at 4°C. Cells were then washed in PBS with 1% BSA, before being permeabilised and fixed according for protocol outlined in eBioScience Foxp3/Transcription Factor Fixation/Permeabilization kit (Invitrogen, catalog: 00-5521-00), before staining with antibodies against IFNy (PE) and TNF (APC) for two hours at room temperature in the dark. Cells were washed in the permeabilisation buffer once before analysis. AIM^+^ cells were identified from non-naïve T cell populations. Gating strategy is outlined in **Supplementary** Figure 3 and **Figure 5A**.

### Primary B cells on CD40Lg and BAFF-expressing feeder cells

To examine the effect of BAFF on BAFF-R in primary B cells from healthy donors, B cells were isolated using magnetic bead separation (Miltenyi Biotec, B Cell Isolation Kit II, catalog: 130-091-151). Following isolation, the B cells were cultured on YK feeder cells engineered to express either CD40Lg, or CD40Lg and BAFF (65). The YK feeder cells, a kind gift from Daniel J. Hodson, were seeded in a 12-well plate at a density of 2 x 10^5^ cells per well in 1 mL of Advanced-RPMI medium (ThermoFisher, catalog: 12633012) supplemented with 20% FBS and 1% GPS (ThermoFisher, catalog: 10378016), and incubated overnight (16-24 h) to allow adherence. Approximately 1 x 10^6^ freshly isolated primary B cells were then added directly on top of the adherent feeder cells, without replacing the existing medium. After 22 hours of co-culture, B cells were examined by flow cytometry as outlined above using the following antibodies: BAFF-R (APC), CD19 (BUV563), CD20 (BUV805) and Zombie NIR Fixable Viability Kit (BioLegend, catalog: 423105).

### Data analysis, statistics, code and visualisation

#### Data analysis

Flow cytometry data was analysed in OMIQ software (www.omiq.ai). For the UMAP, an equal number of B-cell events (CD45^+^CD3^-^CD14C^-^D19^+^ cells) were subsampled (5000 events from each individual) to ensure a fair comparison between individuals. Dataset stained with Spike WT protein was used. In total 48 controls and 22 CVID patients had enough cells and were included. All B-cell markers were used to train the dimension reduction algorithm, and the UMAP calculated using the following settings: neighbors 30, minimum distance 1, components 2, euclidean distance, epochs 200.

Data was subsequently visualised and analysed in either OMIQ, or R programming language version 4.1.1 (66) using custom scripts and RStudio version 2023 (67) 12.0.369 or GraphPad Prism version 10.4.1. The following R packages were used: corrplot 0.92 (68), dplyr 1.0.7 (69), entropy 1.3.1 (70), dendextend (71), Hmisc 4.7.1 (72), lubridate 1.7.10 (73), tidyverse 1.3.1 (74), patchwork 1.1.2 (75), pheatmap 1.0.12 (76), scales 1.1.1 (77).

#### Data availability

Custom R scripts used for calculations used in the paper are available at: https://github.com/mbarnkob/articles/tree/main/2025_Barnkob_BAFFR_CVID, together with further explanation and example data.

#### ED90 decay rate and half-life

The decay rate of neutralizing antibodies over time was calculated by fitting a linear regression model to the data, where the neutralization values were log-transformed with time as the independent variable. The relationship between antibody neutralization and time followed an exponential decay model: y(t) = y0 e^−λt^. By taking the natural logarithm of both sides, we linearized the relationship: ln(y(t)) = ln(y0) − λt. In this linearized equation, the slope of the line (λ) represents the decay rate. The decay rate indicates how quickly the neutralizing ability of antibodies decreased over time. The half-life was calculated: t_1/2_ = ln (2) / λ.

#### Shannon diversity calculations

For analysis of B-cell population diversity, Shannon Diversity was calculated on raw counts of cells in each specified/gated B-cell subpopulation, followed by entropy calculation using maximum likelihood. The Shannon Diversity Index (H’) was calculated using the following formula: H′ = −∑p_i_ ⋅log(p_i_). H’ tells us how evenly distributed the cell counts are across the different subpopulations, with larger values representing a more even distribution across cell clusters and a smaller value representing a less even distribution of cells in subpopulations, e.g. lower cell counts in some cell subpopulations and higher counts in others. Further explanation and example data is provided in the custom R scripts used.

#### Statistics

For the baseline statistics, data are reported as proportions for categorical variables and medians with interquartile range (IQR) for continuous variables. For Pearson r-squared values, all were significant different from 0, unless mentioned. All statistical tests were performed as indicated in figure legends, two-sided with a significance threshold of p < 0.05. * indicates p < 0.05, ** = p < 0.01, *** = p < 0.001, **** = p < 0.0001, and ns indicates not significant. Each dot represents one individual.

#### Visualisation

Figure 1A was created in BioRender under a CC-BY license (https://BioRender.com/d66l499).

## Contributions

Conceptualization: MBB, LDR, TB, CNi, ESL

Methodology: MBB, CNi, OL, LRO, CN, ZU, SJ, RMP, TEA, LDR, DKH

Software: MBB, LRO

Validation: MBB, ZU, ESL, OL, SJ, RMP, TEA

Formal Analysis: MBB, LRO, ZU, KMR, JTS, LDR, TB, RMP, TEA

Investigation: MBB, NB, CLS, ZU, EBC, LLB, DT, ESL, CNi, NDD, OL, RMP, LDR

Resources: MBB, SJ, OL, RMP, TEA, DKH

Data Curation: MBB, CNi, ESL, CLS, NB, LLB, DT, LDR

Writing – Original Draft: MBB, LDR, TB, KMR, LRO, CNi, TEA, ESL

Writing – Review & Editing: MBB, LDR, KM, TB, CNi, ESL

Visualization: MBB

Supervision: TB, MBB, CNi, LDR, SJ, TEA, CN, DKH

Project Administration: LDR, MBB, CNi, ESL, TEA, TB Funding Acquisition: LDR, MBB, CNi, TEA, TB

CNi = Christine Nilsson

## Supporting information

STROBE checkliste

## Data Availability

All data produced in the present study are available upon reasonable request to the authors.

https://github.com/mbarnkob/articles/tree/main/2025_Barnkob_BAFFR_CVID

## Acknowledgements

The authors would like to thank all study participants and donors from OUH Blood Bank for their contribution to this research. We are grateful for technical assistance from personnel from the Leukocyte Laboratory at the Department of Clinical Immunology, OUH, and the research Centre for Cellular Immunotherapy of Haematological Cancer Odense (CITCO), OUH, for helpful discussion and help throughout the project. We wish to thank Professor Daniel J. Hodson for the kind gift of YK6 feeder cells. M.B.B. was supported by Early-Career Clinician Scientists’ fellowship from the Lundbeck Foundation (grant R381-2021-1278) during this work. CITCO is funded through an elite-research grant from Odense University Hospital. The study was supported by The Novo Nordisk Foundation (grant NNF20SA0062931).

## Supplementary information

STROBE reporting guidelines. ICMJE disclosure forms.

Supporting data values.

