## Supplementary Figures for "BAFF-R and CD21 dysregulation inhibits memory B cell persistence in patients with common variable immunodeficiency"

### Supplementary Figure 1

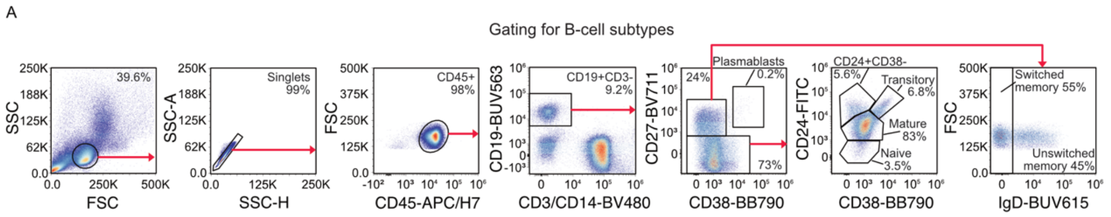


**Supplementary Figure 1.** Gating strategy for identifying B-cell subsets. Related to Figure 3 and 4.

### Supplementary Figure 2

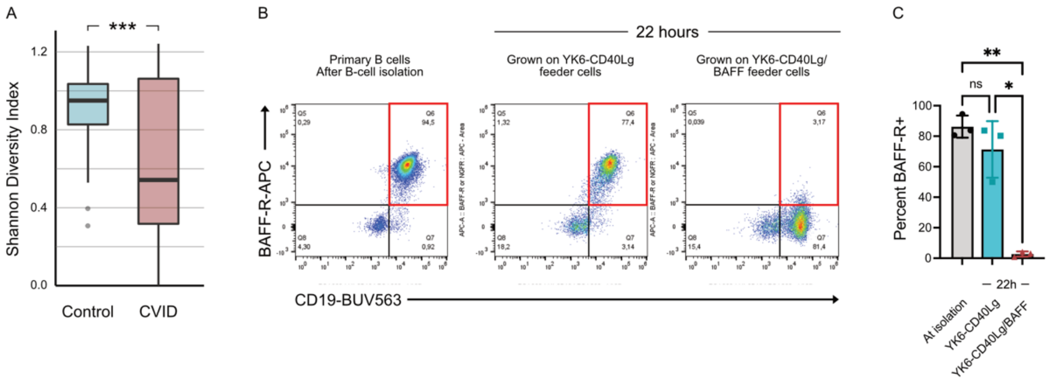


**Supplementary Figure 2. A.** Shannon diversity of B-cell subpopulations in control group and CVID patients. **B**. Analysis of CD19 and BAFF-R on primary B cells from a healthy donor immediately following B-cell isolation, or after 22 hours grown on irradiated YK6 feeder cells genetically engineered to express CD40Lg alone or CD40Lg and BAFF. Red box indicate CD19+BAFF-R+ positive cells. **C.** Quantification of BAFF-R positive cells under same conditions as mentioned in B. Related to Figure 4. Abbreviations: CVID, common variable immunodeficiency disease. * = P < 0.05, ** = P < 0.01, *** = P < 0.001 by Wilcoxon test (**A**) or one-way ANOVA (**C**).

### Supplementary Figure 3

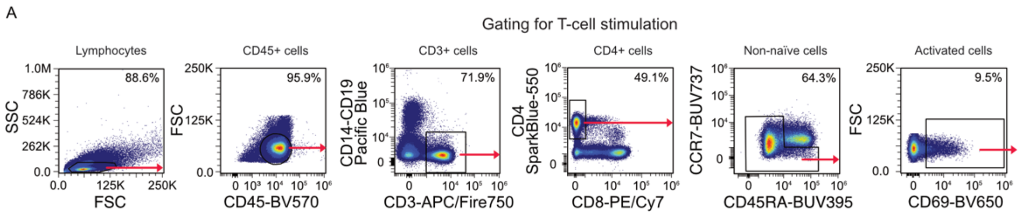


**Supplementary Figure 3.** Gating strategy for identifying non-naïve AIM$^{+}$CD4$^{+}$ T cells following peptide stimulation. Related to figure 5.
